# Serotonin 1A receptor distribution in treatment-resistant depression during psychopharmacotherapy compared to healthy controls

**DOI:** 10.1101/2023.09.29.23296333

**Authors:** Matej Murgaš, Christian Milz, Peter Stöhrmann, Godber M. Godbersen, Jakob Unterholzner, Lukas Nics, Georg S. Kranz, Andreas Hahn, Marcus Hacker, Siegfried Kasper, Rupert Lanzenberger

## Abstract

Major depressive disorder (MDD) is associated with a high lifetime prevalence and is a major cause of disability. An additional burden on patients is that up to 60% of the first antidepressant (AD) trials do not provide adequate symptom relief and after two subsequent AD trials, a patient is referred to as treatment-resistant. The serotonin 1A receptor subtype (5-HT_1A_) has commonly been used to study pathophysiological alteration in MDD. However, PET data on treatment-resistant depression (TRD) populations is still limited.

In this cross-sectional study, 5-HT_1A_ receptor binding was assessed in 20 TRD patients (9 female, mean age ± SD, 29.0 ± 5.2) and 20 healthy controls (HC) (10 female, mean age ± SD, 33.2 ± 8.2). Positron emission tomography (PET) scans with the radiotracer [*carbonyl*-^11^C]WAY-100635 were acquired and 5-HT_1A_ non-displaceable binding potential (BP_ND_) was quantified using the multilinear reference tissue model 2, with the cerebellar white matter as reference region. Mean regional BP_ND_ in five regions of interest (amygdala, anterior cingulate cortex, hippocampus, insula and orbitofrontal cortex) was compared in a repeated measures analysis of covariance (rmANCOVA) with age, sex and group as covariates.

Estimated marginal means showed slightly lower BP_ND_ in TRD group (mean ± SD = 5.464 ± 0.247) than in the HC group (mean ± SD = 5.938 ± 0.245). However, the rmANCOVA showed no significant group difference (p = 0.659).

Studies on 5-HT_1A_ binding in MDD show heterogeneous results, where the directionality of difference as well as the significance of findings strongly depend on specific outcome measures (BP_ND_, BP_F_ or BP_P_), reference region or quantification method. Here we showed no significant effect of TRD on BP_ND_, similar to other studies applying the same methodology for MDD cohorts.

## INTRODUCTION

Major depressive disorder (MDD) exhibits high lifetime prevalence and is one of the most common psychiatric disorders. In addition to being associated with a multitude of chronic disorders (e.g. asthma or hypertension), it also typically affects the financial and social aspects of life (1,2) and is connected to increased mortality (3). Despite having access to a variety of antidepressant treatment possibilities, only 40% of patients respond to first-line pharmacological treatment. A subpopulation of these non-responders even fail to respond to two adequate antidepressant trials (i.e. psychotherapeutic and/or pharmacological), thus representing treatment-resistant depression (TRD) patients (4). This could be due to a host of risk factors including higher suicidal risk or higher comorbid anxiety disorder (5).

The involvement of the serotonergic system in major depressive disorder has been extensively investigated in past decades (6). In this context, positron emission tomography (PET) has become a main modality for investigating pathophysiological states in psychiatric disorders *in vivo*. Previous studies involving the serotonergic system showed alterations in MDD subjects. For example, longitudinal effects SSRI treatment in MDD subjects have been shown measuring SERT occupancy before and after treatment (7). Another study unveiled that changes to 5-HT synthesis contributed to the progress of MDD (8). In particular, the serotonin 1A (5-HT_1A_) receptor is one of the most abundant receptors of the serotonergic system in the human brain, and thus also the most researched subtype of serotonin receptors. In the past, pathophysiological states and changes to its distribution in the cortex were connected to different psychiatric disorders, including depression (9). For instance, the reduction in 5-HT_1A_ receptor availability across different brain regions was demonstrated in various cohorts of MDD patients (10,11). Specifically, receptor-rich regions of amygdala, anterior cingulate cortex, hippocampus, insula and orbitofrontal cortex, showed a decrease in several studies (12–14). Additionally, Albert et al. (11) were able to show that altered 5-HT_1A_ receptor transcription affects the serotonergic system and might lead to depression.

In addition to insufficient or no response to conventional antidepressant treatment, several risk factors have been identified for TRD patients (5). Therefore, treatment-resistant depression should be seen as a condition extending beyond MDD. As such, investigations of 5HT_1A_ receptor distribution, although extensively researched in MDD, are needed to investigate possible alterations in TRD. Until now, investigations on treatment-resistant depression included genetic factors (15) but assessment of 5HT_1A_ receptor binding is limited.

In this work, we aim to unveil the differences in 5-HT_1A_ binding between treatment-resistant depressive patients and healthy control subjects. In particular, we aim to investigate the differences in the high binding regions using *in vivo* positron emission imaging with the specific radioligand [*carbonyl*-^11^C]WAY-100635. In line with previous extensive work on major depression and associated changes to the serotonergic system, we hypothesize that there is a significant decrease in serotonin 1A receptor binding potential in TRD patients.

## MATERIAL and METHODS

### Subjects and study design

The dataset represents a collection of published studies alongside previously unpublished data. The data of healthy control subjects (HC) were taken from Lanzenberger et al. (16) and Baldinger et al. (17) (Registered at International Standard Randomized Controlled Trial Number Register as ISRCTN30885829). The part of treatment-resistant depression patient group data were adopted from Murgaš and Unterholzner et al. (18) and complemented by an additional set of unpublished data collected from the same study (Registered at ClinicalTrials.gov as NCT02810717). For details on demographic information, see the Table 1 in results section. Only baseline PET measurements of the patients were included. However, patients were allowed for concomitant medication if it was stable for at least 4 weeks prior to the recruitment to study. To screen subjects for physical abnormalities, routine medical examinations were performed, including laboratory measurements, general physical and neurological status and an electrocardiogram. Moreover, clinical interviews by experienced psychiatrists ensured mental health of the HC group and excluded psychiatric comorbidities in TRD patients. The TRD group was recruited from the outpatient department and the hospital wards of the Department of Psychiatry and Psychotherapy at the Medical University of Vienna, Austria. Each patient was carefully screened using SCID IV (Structural Clinical Interview for DSM IV Diagnosis) by trained physician and included in the study only when fulfilling the criteria for single or recurrent major depression and having a score ≥ 18 on 17-item Hamilton Rating Scale for Depression (HAMD). Subjects gave informed written consent at the screening visit. All procedures were performed according to Declaration of Helsinki. All studies were approved by the Ethics Committee at the Medical University of Vienna (318/2002; 475/2011; 1761/2015).

**Table 1:** Demographic information on treatment-resistant depressive (TRD) patients and healthy control (HC) subjects. F=Female, M=Male, HAMD=17-item Hamilton Rating Scale for Depression.

|  | TRD patients | HC subjects |
| --- | --- | --- |
| N | 20 | 20 |
| Sex F/M | 9/11 | 10/10 |
| Age (mean ± SD) | 29.0 ± 5.2 | 33.2 ± 8.2 |
| HAM-D 17 (mean ± SD) | 21.3 ± 3.6 | N/A |

### Neuroimaging

Imaging procedures for HC and TRD groups were similar although details may vary. For each subject, a 90-minute PET scan was acquired, using a GE Advance PET scanner (General Electric Medical Systems, Milwaukee, Wisconsin) at the Department of Biomedical Imaging and Image-guided Therapy, Division of Nuclear Medicine, Medical University of Vienna, Austria. Acquisition in 3-D mode started with bolus infusion of the radioligand [*carbonyl*-^11^C]WAY-100635 with a dose of 4.6-5.4 MBq/kg. A 5-minute transmission scan in 2-dimensional mode with a retractable ^68^Ge source was used for attenuation correction. Images were reconstructed as a 128 × 128 matrix (35 slices) with an iterative filtered back-projection algorithm with a spatial resolution of 4.36 mm full-width at half-maximum 1 cm next to the center of the field of view. Here, the number and the length of frames in the reconstructed dynamic images varies across the studies: Lanzenberger et al. uses 30 time frames (15 × 60 sec and 15 × 300 min), Baldinger et al. uses 50 time frames (12 × 5 sec, 6 × 10 sec, 3 × 20 sec, 6 × 30 sec, 4 × 60 sec, 5 × 120 sec and 14 × 300 sec) and patients group is reconstructed to 51 frames (12 × 5 sec, 6 × 10 sec, 3 × 20 sec, 6 × 30 sec, 9 × 60 sec, and 15 × 300 sec). The radioligand [*carbonyl*-^11^C]WAY-100635 was prepared at the cyclotron unit of the PET Center according to previously published methods (19).

Additionally, a structural T1-weighted MR image was acquired for each subject. TRD subjects were scanned at a 3T PRISMA MR Scanner (Siemens Medical, Erlangen, Germany) using the magnetization-prepared rapid gradient-echo sequence (TE/TR = 4.21/3000 ms, voxel size 1 × 1 x1.1 mm^3^).

### Data processing and quantification

Each PET scan was corrected for attenuation and scatter. Preprocessing was performed using SPM12 (The Wellcome Centre for Neuroimaging, www.fil.ion.ucl.ac.uk) and MATLAB R2018b (The Mathworks Inc., Natick, MA, USA). Correction for head motion was applied followed by its co-registration to the T1-weighted structural MR image. The structural image was then normalized to Montreal Neurologic Institute (MNI) space and the same transformations were applied to the co-registered dynamic PET images as well.

Time activity curves were extracted from the normalized PET scans for insula and cerebellar white matter (CWM). The former shows a high specific uptake of [*carbonyl*-^11^C]WAY-100635 whereas CWM is devoid of specific binding, making it a suitable choice for reference tissue models (20). Nondisplaceable binding potential (BP_ND_) was modelled with the multilinear reference tissue model 2 (MRTM2) (21) using the TACs as input in PMOD, version 3.5 (PMOD Technologies LLC, https://www.pmod.com/web/). Regions of interest were selected according to published data on changes in 5-HT_1A_ binding in amygdala (AMY), anterior cingulate cortex (ACC), hippocampus (HIP), orbitofrontal cortex (OFC) and insula (INS) in patients suffering from major depression disorder (22). Mean BP_ND_ was extracted for each ROI using an in-house atlas (23).

### Statistical analysis

To correct for the different study protocols (i.e., frame length) of the PET scans included in the healthy control group, data harmonization was applied. Here we used the Matlab implementation of the ComBat toolbox (24,25) with age and sex as a covariate.

To test for the differences in 5-HT_1A_ receptor distribution between healthy control subjects and patients we used a repeated measures analysis of covariance (rmANCOVA). Selected regions of interest were used as repeated measures, while sex and subject groups as factors and age as covariate. If not stated otherwise, all statistical analysis were performed in SPSS version 26 for Windows (SPSS Inc., Chicago, Illinois, USA; www.spss.com).

## RESULTS

Data from 40 subjects were available for the comparison of serotonin 1A receptor distribution in selected regions. In particular, 20 HC subjects (mean age ± SD, 33.2 ± 8.2, 10 female) and 20 TRD patients (mean age ± SD, 29.0 ± 5.2, 9 female) were analyzed for this study.

Average regional BP_ND_ values of amygdala, anterior cingulate cortex, hippocampus, insula and orbitofrontal cortex showed lower binding in TRD patients than in HC subjects (see Figure 1).

**Figure 1:**
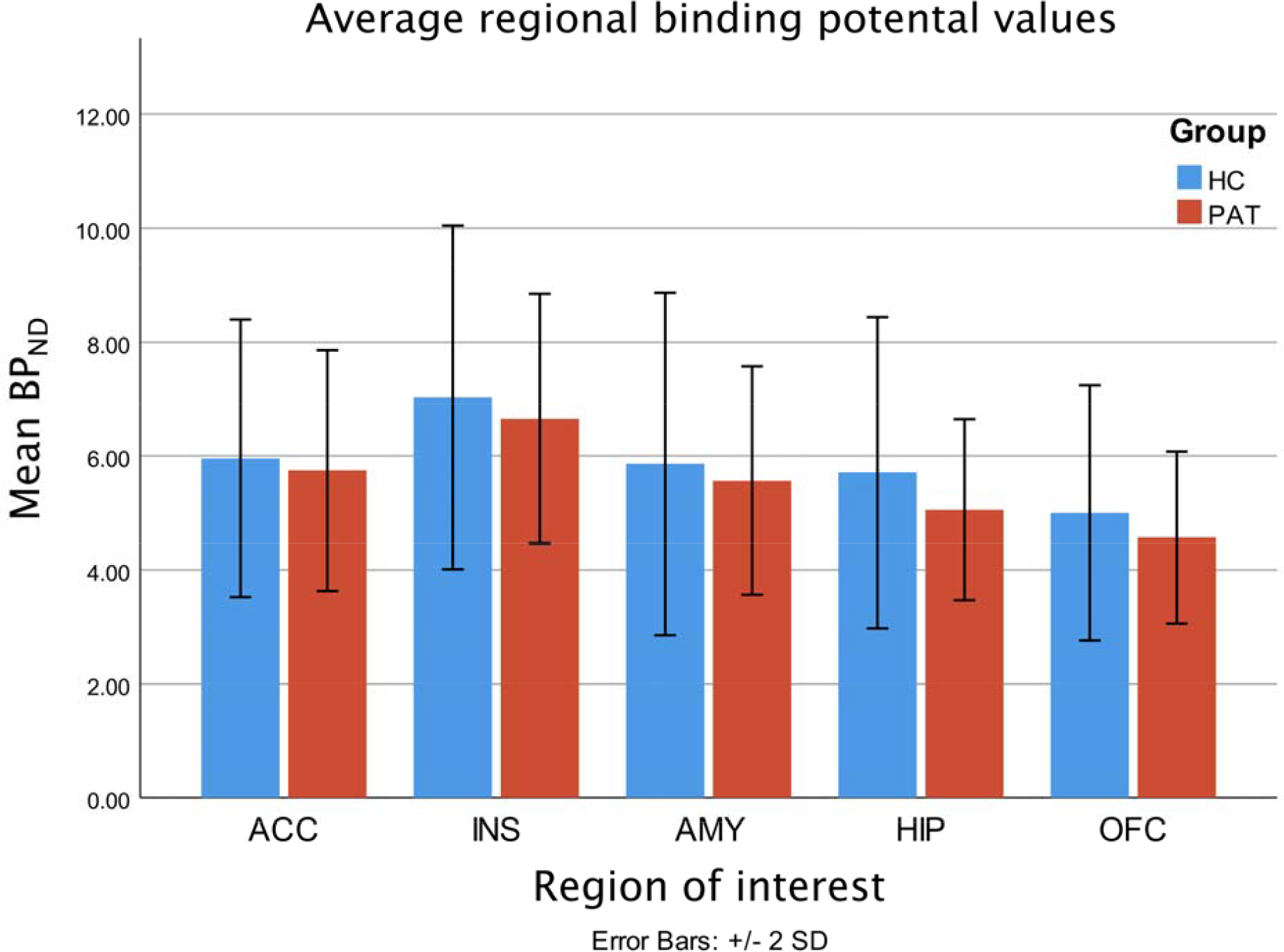
Barplot showing average BP_ND_ values of amygdala (AMY), anterior cingulate cortex (ACC), hippocampus (HIP), insula (INS) and orbitofrontal cortex (OFC) in both subject groups of healthy control subjects (HC) and TRD patients (PAT).

The rmANCOVA, using regional BP_ND_ values as repeated measures, showed no significant between-subject effect of the group (F = 1.747, p = 0.195), sex (F = 0.022, p = 0.882), age (F = 0.198, p = 0.659) nor a two-way interaction between sex and group (F = 3.617, p = 0.065). Despite the non-significant result, the estimated marginal means showed slightly lower BP_ND_ in the TRD group (mean ± SD = 5.464 ± 0.247) than in HC group (mean ± SD = 5.938 ± 0.245). The low numerical difference between the groups is supported also by the visual comparison of 5-HT_1A_ distribution (see Figure 2).

**Figure 2:**
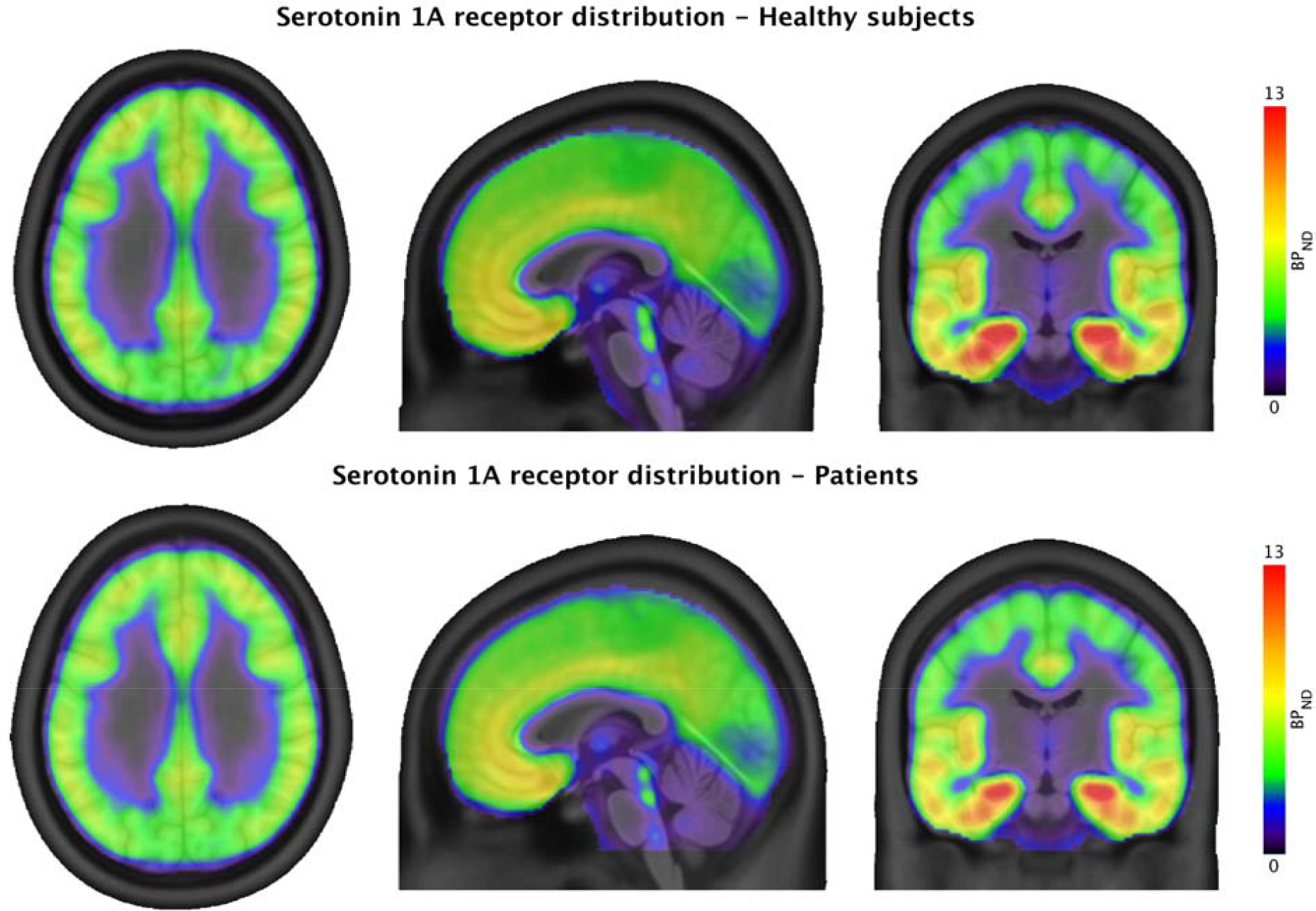
The tri-planar visualization (MNI coordinates x = 0, y = -14, z = 30) of serotonin 1A receptor distribution measured by PET with [carbonyl-^11^C]WAY-100635 radioligand in healthy subjects group (top row) and TRD patients group (bottom row).

## DISCUSSION

In this study, we investigated the differences between treatment-resistant depressive patients and healthy control subjects in the 5-HT_1A_ receptor binding potential. Despite minor visual differences, we could not show a significant difference in serotonin 1A receptor binding between TRD and HC subjects across the investigated regions. In line with our results, Parsey et al. (13), using simplified reference tissue modeling (SRTM) with the CWM as a reference region, were not able to show a significant change in 5-HT_1A_ BP_ND_ in MDD patients. On the other hand, other study showed a non-significant increase in remittent MDD patients in comparison to HC subjects using SRTM with CWM as a reference region (26).

In contrast to the other studies, where MDD patients are typically drug-naïve, here we investigate a specific subgroup of treatment-resistant depressive patients who are medicated. Each patient was under medication affecting the serotonergic system and underwent at least two different pharmacological treatments in the past. Previous research has showed that such treatment might have region and substance-specific effects (27). In addition, long-term pharmacological treatment was suggested to reduce 5-HT_1A_ concentrations in limbic regions (28) as well as in hippocampus (29). On the other hand comparing antidepressant medicated and drug-naïve patients showed no differences in BP_ND_ between these two groups (30,31). Hence even though the possible effect of the treatment cannot be ruled out, we expect it could be negligible. Therefore our results in TRD patients can be compared to existing studies with MDD patients.

Publications with similar research aims and sample size using a MDD cohort but with a different methodology, report varying results, ranging from reduced 5-HT_1A_ receptor binding to a lack of significant differences or even increased binding. Hence, a consensus has been reached that the apparent contrasting effects of MDD on serotonin 1A receptor binding can be by large accredited to methodological discrepancies i.e. compared output measure (32). Multiple studies using the optimal modeling method of the two-tissue compartment model (2-TCM) with plasma-corrected arterial input function (AIF) have shown significant changes in 5-HT_1A_ binding. While BP_F_ was shown to increase in MDD patients (13,33), Hirvonen et al. (14) showed a significant decrease in BP_P_. On the other hand, no changes to BP_ND_ in MDD patients were shown using 2-TCM with AIF (13,14). Inconsistency of the results based on the output measure emphasizes the role of careful interpretation of the results of the studies on 5-HT_1A_ receptor binding changes in depressive patients.

The validity of the cerebellum or parts of the cerebellum as reference region for quantification of [*carbonyl*-^11^C]WAY-100635 binding has been extensively discussed. Comparison of the quantification results using different parts of the cerebellum as a reference region to the gold standard AIF modeling across multiple brain regions has also confirmed non-optimal performance of cerebellar gray matter (CGM) as a reference region while suggesting CWM as a better option (34). Higher 5-HT_1A_ receptor abundance was suggested in blocking experiment that has showed stable levels of volume of distribution in CWM and decreased values of volume of distribution in CGM following pindolol application (13). This finding is also confirmed by autoradiogram results showing the lowest 5-HT_1A_ receptor concentration in cerebellar white matter (20). Thus, results using other than cerebellar white matter as reference region should be interpreted with caution.

Some limiting factors to the study design have been identified. First, the control group comprises subjects from two different study setups. Having HC subjects and TRD patients from the same study would reduce differences in image acquisition and processing. However, differences within the HC group are only related to the reconstructed frame length of PET scans, which were accounted for by applying data harmonization applied after the BPND quantification. Second, the lack of arterial blood samples prevented us from comparing BP_ND_ to alternative outcome measures such as BP_P_ or BP_F_ using the gold standard of 2-TCM plus AIF. Finally, although patients did not change the medication, the diversity of the accompanying treatment might affect the final result. However due to the sample size and variety of concomitant medication such effect could not be investigated in the current study.

In conclusion, we were not able to show a significant decrease in serotonin 1A receptor concentration in treatment-resistant depressive subjects. This result is in line with previous findings with respect to the used methodology.

## Data Availability

Raw data will not be publicly available due to reasons of data protection. Processed data and custom code can be obtained from the corresponding author with a data-sharing agreement, approved by the departments of legal affairs and data clearing of the Medical University of Vienna.

## ACKNOWLEDGEMENTS

This research was funded in whole, or in part, by Austrian Science Funds (FWF) [Grant number KLI 1006 to R. Lanzenberger, KLI 551 to S. Kasper]. M. Murgaš was funded by the Austrian Science Fund (FWF) [Grant number DOC 33-B27, Supervisor R. Lanzenberger].

We would like to express our gratitude towards Pia Baldinger-Melich, Gregor Gryglewski, Marius Hienert, Marie Spies, Christoph Kraus, Alexander Kautzky, Arkadiusz Komorowski, Paul Michenthaler, and Richard Frey for clinical support, and Murray B. Reed, Lucas Rischka and Sebastian Ganger for technical support, and all additional staff and students from the Neuroimaging Lab (NIL) involved in the realization of this research. We would like to thank to Department of Biomedical Imaging and Image-guided Therapy, Division of Nuclear Medicine, Medical University of Vienna, especially Cécile Philippe, Chrysoula Vraka and Wolfgang Wadsak, for support in radiosyntheses, close cooperation and technical support. Moreover, we would like to thank radiotechnologists, Ingrid Leitinger and Harald Ibeschitz, for operating PET.

